# Prevalence and Risk Factors of Stroke in Bangladesh: A Nationwide Population-Based Survey

**DOI:** 10.1101/2021.06.17.21259097

**Authors:** Md Badrul Alam Mondal, A T M Hasibul Hasan, Nushrat Khan, Quazi Deen Mohammad

**Affiliations:** National Institute of Neurosciences and Hospital; University of Cambridge, UK

## Abstract

**Background:** This community survey was conducted to estimate the prevalence of stroke and its associated common risk factors among the Bangladeshi population.

**Methods:** This was a population-based cross-sectional study, carried out in 8 administrative divisions and 64 districts to estimate the prevalence of stroke throughout the country. The study adopted a two-stage cluster random sampling approach. The calculated sample size was 25,287. A semi-structured questionnaire was used to identify suspected stroke patients who were subsequently confirmed by consultant neurologists.

**Result:** In the first stage, Interviewers identified 561 respondents as suspected stroke in 64 districts. Of the 25,287 respondents, 54.9% were male. The mean age was 39.9 years. In the second stage of the study; among all the respondents, 288 were confirmed as stroke patients which provided a prevalence of 11.39 per 1000 population. The highest stroke prevalence (14.71 per thousand) was found in the Mymensingh division and the lowest (7.62 per thousand) found in the Rajshahi division. It was 30.10 per thousand in the age group of more than 60 years. The prevalence of stroke among males was twice that of females (13.62 versus 8.68 per thousand). The prevalence was slightly higher in rural areas (11.85 versus 11.07). Out of a total of 288 cases, 79.7% (213) patients had an ischemic stroke, 15.7% (42) had hemorrhagic, and 4.6% (12) were diagnosed as subarachnoid hemorrhage. The majority of the stroke patients had hypertension (79.2%), followed by dyslipidemia (38.9%), tobacco use in any form (37.2%), diabetes (28.8%), ischemic heart disease (20.1%).

**Conclusion:** We have found a stroke prevalence of 11.39 per 1000 population, the highest being in the Mymensingh division. The prevalence was much higher in the elderly and male population. More than three fourth had an ischemic stroke. Hypertension, dyslipidemia, tobacco use, diabetes, ischemic heart disease are the most common risk factors observed among stroke patients.

**Summary Box:** Already Known:

▪ The prevalence of stroke in Bangladesh was found to be 3 per thousand.
▪ Hypertension, diabetes, dyslipidemia, smoking, and ischemic heart disease were common risk factors.

New Findings:

▪ This is the first-ever nationwide survey in Bangladesh that revealed a stroke prevalence of 11.39 per thousand.
▪ There was a wide regional variation of stroke prevalence.
▪ The prevalence was twice among males.

Impact of the study result:

▪ The study result will help the policymakers in deciding what to do for which regions of this country to handle the stroke burden.
▪ It will also help the clinicians to identify common risk factors among stroke patients.

## Introduction

A major a socio-demographic and economic shift has been observed in Bangladesh over the past few decades. Bangladesh has recently joined the list of lower-middle-income countries and expecting progress to the grade of a middle-income country by 2026 The relationship between economic growth and non-communicable diseases (NCDs) is complex. With this dynamic economy and achieving the targets of millennium development goal, Bangladesh observed an increase in life expectancy and therefore, a growing number of the aging population. Sustainable economic growth is impossible without addressing NCDs as it is a major cause of disability leading to health system burden^1^.

Around 80% of NCD death occurs in low and middle-income countries (LMICs). It is evident amongst NCDs, stroke is a major cause of death and disability globally. Stroke devastates life with 10.3 million new cases per year, 6.5 million deaths, and around 26 million survivors with some disability^2^. The incidence of stroke in LMICs has increased from 56/100,000 person-years to 117/100,000 person-years throughout three decades^3^. More than half of annual deaths due to NCDs are reported from South-East Asian countries^3^. Considering the poor disease reporting or death registration systems in LMICs, the epidemiological findings from the Global burden of Disease study for many of the LMICs might not be a reflection of the actual burden.

With this fast economic transition, the burden of diseases in Bangladesh is shifting from infectious disease to non-communicable ones. With epidemiological transition, the major cause of death has gradually shifted from infectious and parasitic diseases to NCDs over the last three decades^4^. Limited available data suggest that in Bangladesh around 51% death occurs due to NCDs^4^. Bangladesh is ranked ninth among the countries with the highest degree of age-standardized mortality owed to chronic disease, primarily cardiovascular disease and diabetes^5^.

On the contrary to the disease burden, there is a scarcity of data on stroke epidemiology in Bangladesh. Two community level surveys were conducted previously on the prevalence of stroke in Bangladesh which involved only a small area^6,7^. Considering the epidemiologic shift and weak public health reporting mechanism; making a judicious policy to mitigate the disease burden is a mammoth task in Bangladesh given the low healthcare expenditure.

Therefore, we conducted this nationwide stroke survey to gather evidence on the current prevalence and risk factors of stroke among Bangladeshi population.

## Methodology

We had conducted this population-based cross-sectional study, throughout 8 administrative divisions and 64 districts to estimate the prevalence of stroke in the country during a period of six months from January 2018 to June 2018, involving a representative sample of adults aged 18 years and above residing anywhere in Bangladesh. The study adopted a two-stage cluster random sampling approach. The selected districts were the Primary Sampling Units (PSUs) while the targeted respondents were the final sampling unit (FSU) for the survey. The allotted numbers of clusters in each division were proportionate to the population size of that division (Supplementary Table -1). The number of PSUs selected in each division was equal to the allotted number of clusters. Then, each randomly chosen ward or mauzas had been divided arbitrarily into segments. The sample size was determined considering the anticipated population prevalence, precision rate, confidence interval of 95%, 80% power and design effect. The calculated sample size was 25,278. So, 180 participants in one segment were finalised by dividing the total sample size 25,278 by 140 clusters. The number of segments of each ward or village was determined by dividing the total adult population of the ward or village by the cluster size of 180. Then one segment containing 180 people from each randomly selected ward or village was again selected randomly for the interview. If a segment did not cover a required number of households, (which was unlikely) then the households meeting inclusion criteria from the adjacent segment/village were taken to complete the target. The total number of segments considered to interview samples equal to the total number of clusters.

The inclusion criteria for this study were consenting males and females aged 18 years and above living in the community, having hemiplegia/ monoplegia/ difficulty in speech and swallowing. Patients with transient ischaemic stroke (TIA), head injury, brain tumour, and other stroke mimics were excluded from the study with documented verification of the diagnosis. Physical disability or serious physical illness was also an exclusion criteria for this study. Written informed consent was taken from all the participants. We formulated 25 teams of data collectors. Each team consisted of 2 members and covered mostly three administrative districts. Each of them received 7 days of intensive training by the researches/medical doctors. Then 2 days of field testing was performed on 100 randomly selected individuals before finalising the questionnaire.

In the first stage of data collection, the data collectors used a household questionnaire where the demographic characteristics and general stroke-related questions of respondents were asked. In later stage, the validated Questionnaire for Verifying Stroke Free Status (QVSFS) was used to screen for stroke patients. Researchers supervised and randomly checked questionnaires throughout.

Proper, encoding, cleaning, and processing of data were done before analysis. Data analysis was done by statistical package for social sciences (SPSS) version 21.0. Descriptive and inferential statistics were used for data analysis. Relevant data were presented by using various diagram and charts.

### Ethical approval

Before the commencement of this study, the research protocol was approved by the Institutional Review Board of the National Institute of Neurosciences and Hospital.

## Patient and Public Involvement

We conducted stroke clinic at our institute thrice a week where we got some rough idea about the basic demographic distribution of our stroke patients. We had a informal discussions with patients and their attendants at the clinic about the probable stroke cases in their vicinity and the distribution of risk factors. With this background knowledge, we had conducted two small-scale surveys in two different regions of Bangladesh (Gazipur and Shirajganj) that involved only a single administrative area which yielded stroke a prevalence of around 3 per thousand. Then we also discussed with patients randomly about conducting a nationwide study and the probable risk factors in our population which will help in further policymaking and health service delivery to our stroke patients. They were assured that the result of the study will be disseminated by national media.

## Results

### Demographic profile

The research team interviewed a total of 25287 respondents. About one-third of the respondents (7577, 30%) were from the Dhaka division, followed by Chittagong (4801, 19%), Rajshahi (3282, 13%), Rangpur (2789, 11%), Khulna (2783, 11%), Sylhet (1767, 7%), Barisal (1268, 5%), and lowest from Mymensingh division (1020, 4%) (Figure-I).

A majority (59.4%) of the respondents in this study were in younger age range (<40 years), followed by 19.6% were in the 41-50 years group, 12.3% in the 51-60 years group and 8.7% were in more than 60 years. The age range was 18 to 113 years. The mean age of the respondents was 39.97 ± 14.03 years. Just about half (13878, 54.9%) of the respondents were male and the rest (11409, 45.1%) were female with a male to female ratio of 1.21: 1 (Table-1). Around 58.9% (14904) of the total respondents were from the urban areas and the rest (41.1%) were from rural areas (Table-1).

**Table 1:** Distribution of the respondents by age (N= 25287).

| Age Group (years) | Male<br>13878(54.9) | Female<br>11409(45.1) | N (%) |
| --- | --- | --- | --- |
| 18 -30 | 3734 (26.9) | 3857 (33.8) | 7591 (30.0) |
| 31-40 | 3644 (26.3) | 3405 (29.8) | 7049 (27.9) |
| 41-50 | 2831 (20.3) | 2152 (18.9) | 4965(19.6) |
| 51-60 | 1987 (14.3) | 1132 (9.9) | 3119(12.3) |
| >60 | 188 (1.4) | 182 (1.4) | 370 (1.5) |
| **Residence** |  |  |  |
| Urban | 7983 (57.6) | 6921 (60.6) | 14904(58.9) |
| Rural | 5887 (42.4) | 4496 (39.4) | 10383(41.1) |
| **Marital status** |  |  |  |
| Married | 11803 (85.1) | 10044 (88.0) | 21847 (86.4) |
| Single | 1956 (14.1) | 510 (4.5) | 2466 (9.8) |
| Divorced/widowed | 111 (0.8) | 863 (7.6) | 974 (3.9) |
| Education level |  |  |  |
| No education | 2922 (21.1) | 3043 (26.7) | 5965 (23.6) |
| Under 10<sup>th</sup> | 5159 (37.2) | 5268 (46.1) | 10427 (41.2) |
| SSC pass | 2230 (16.1) | 1656 (14.5) | 3886 (15.4) |
| HSC pass | 1653 (11.9) | 843 (7.4) | 2496 (9.9) |
| Graduate | 1277 (9.2) | 406 (3.6) | 1683 (6.7) |
| Postgraduate | 629 (4.5) | 201 (1.8) | 830 (3.3) |

**\*\*\*all the variables are statistically significantly different in between gender**

### Prevalence of stroke

The survey revealed out of 25287 respondents, 288 were diagnosed with a stroke at least once in lifetime providing prevalence of 11.4 per 1000 population. The highest stroke prevalence was found in the Mymensingh division (14.71 per thousand), followed by Khulna (14.01), Barishal (13.41), Dhaka (12.27), Sylhet (11.88), Chattagram (11.04), Rangpur (8.96), and the lowest was 7.62 per thousand in Rajshahi division (Table-2).

**Table-2:** Prevalence of stroke by division

| **Division** | **Stroke Cases** | **Respondents** | **Prevalence per 1000** |
| --- | --- | --- | --- |
| Rajshahi | 25 | 3282 | 7.62 |
| Khulna | 39 | 2783 | 14.01 |
| Barisal | 17 | 1268 | 13.41 |
| Dhaka | 93 | 7577 | 12.27 |
| Mymensingh | 15 | 1020 | 14.71 |
| Chattagram | 53 | 4801 | 11.04 |
| Sylhet | 21 | 1767 | 11.88 |
| Rangpur | 25 | 2789 | 8.96 |
| **Total** | **288** | **25287** | **11.39** |

The stroke prevalence was highest (30.10 per thousand) among people aged &#x2265; 60 years and lowest in the age group of 18 to 40 years (4.60 per thousand). The prevalence was 14.7 per thousand in the age group of 41-50 years and 25.2 per thousand in the age group of 51-60 years (Table-3). The prevalence of stroke among males was 13.8per thousand and among females 8.68 per thousand (nearly twice that of females). While the prevalence of stroke in rural areas was 11.85 per thousand, it was 11.07 per thousand in the urban area (Table-3).

**Table-3:**
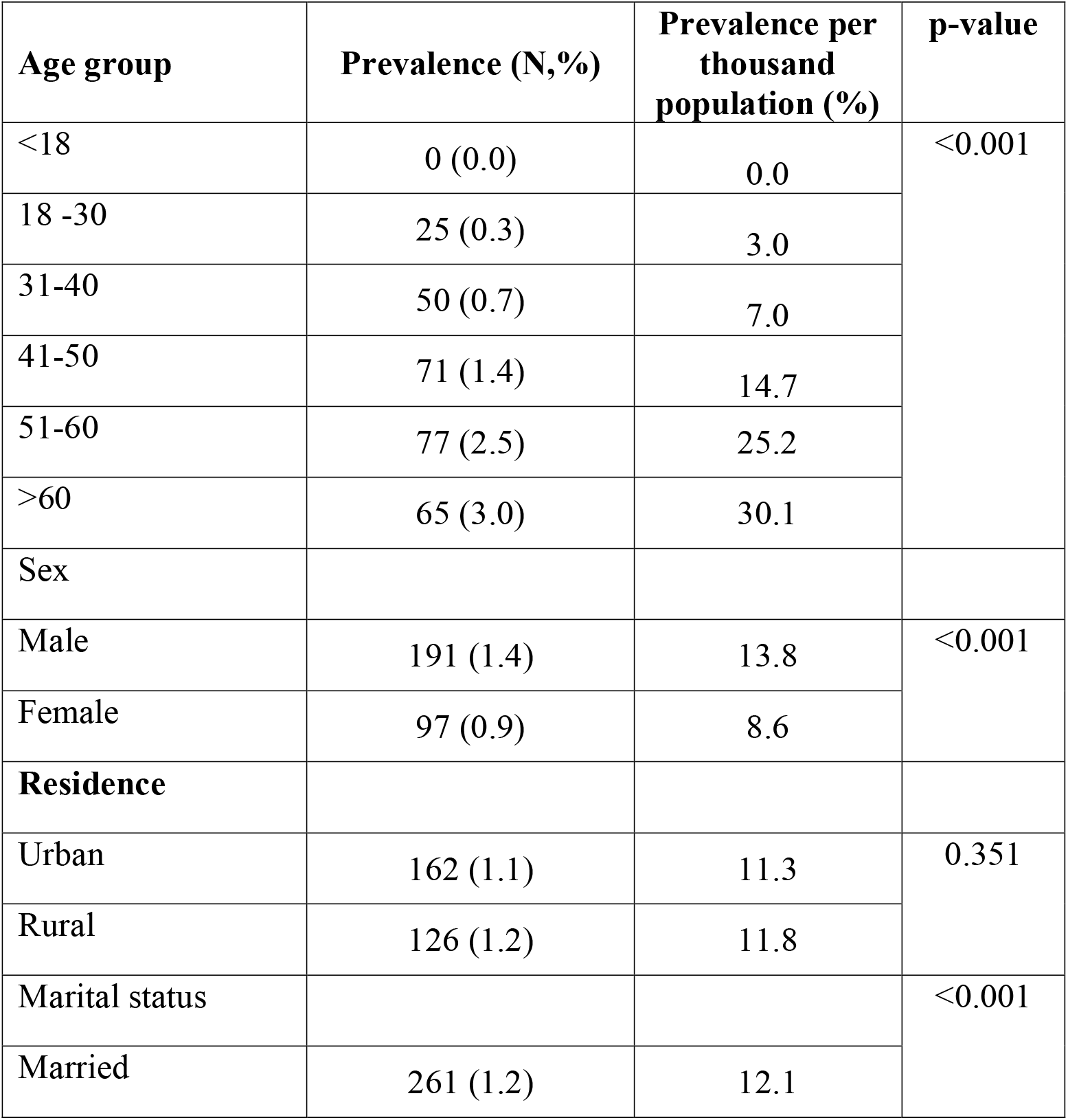
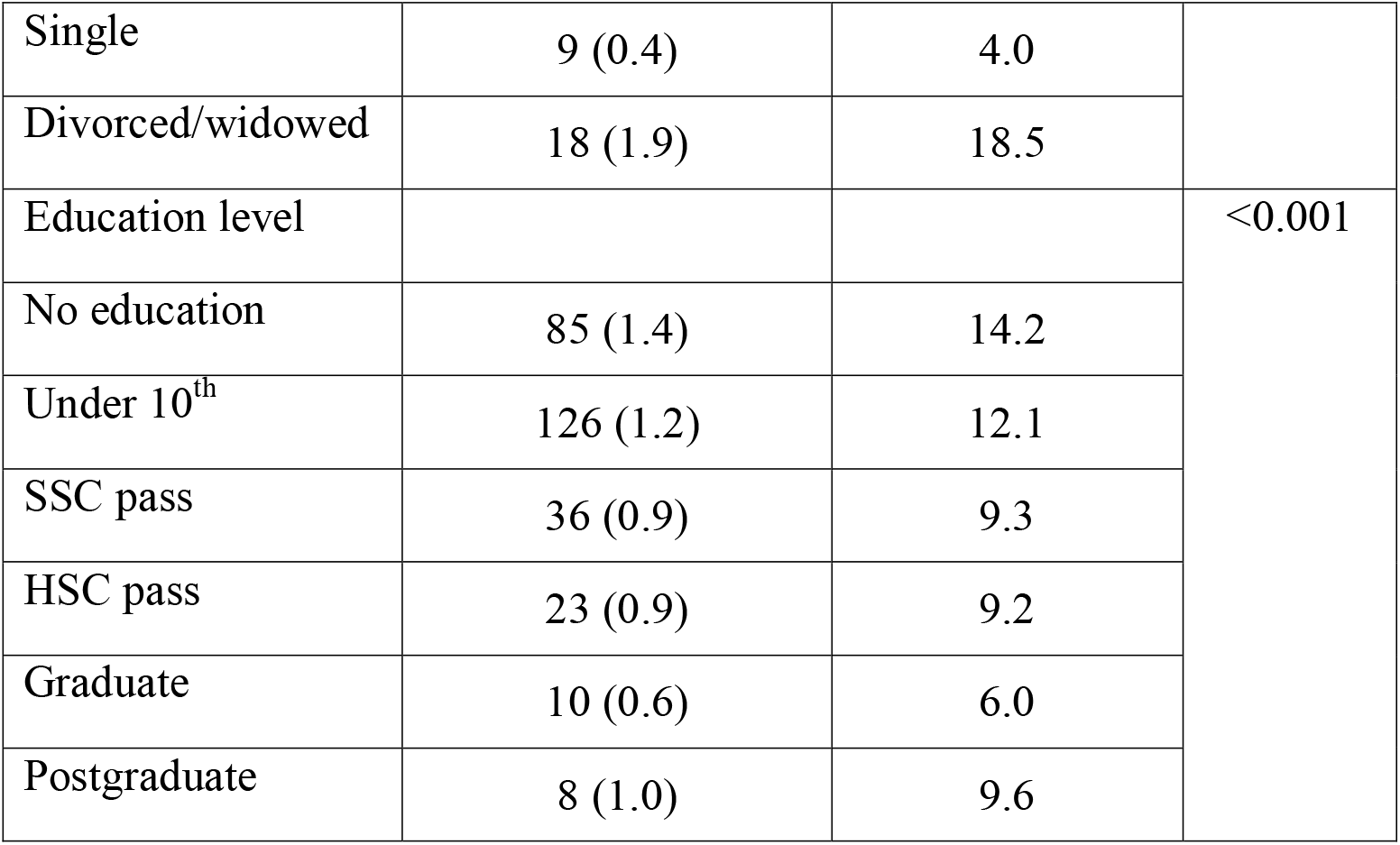
Age, sex, and residence-specific prevalence of stroke.

### Types and risk factors

Out of 288 stroke cases, the majority had an ischemic stroke (213, 79.7%), followed by intracerebral hemorrhage (42, 15.7%) and subarachnoid hemorrhage (12, 4.6%) (Figure-II).

In table 4, we can see the distribution of stroke patients in terms of age, BMI and waist hip ratio. Mean age of the stroke patients are significantly higher then patients without stroke. Amongst male and female, male participents had higher BMI than female which is significantly different. Through the questionnaire, we have tried to outline the distribution of common risk factors of stroke among the patients. Hypertension was the most common risk factor observed among the stroke patients (79.2%), followed by the dietary habit of lack of consumption of recommended daily servings of fruits and vegetables (67.3%). Dyslipidemia and smoking including tobacco use were the next common entity (38.9% and 37.2%) in stroke patients followed by Diabetes (28.8%). History of ischemic heart disease (IHD) was present in 20.1% of stroke patients, atrial fibrillation in 13.9%, and valvular heart disease only in 1% of patients. About 9.7% of patients were obese (Figure-III).

**Table 4:** Subgroup analysis for stroke patients (for body measurements):

| Characteristics | Male | Female | Overall |
| --- | --- | --- | --- |
| Age (mean, SD) | 53.8 (13.4) | 49.6 (12.3) | 52.3 (13.1) |
| BMI | 24.0 (4.6) | 23.8 (4.1) | 23.9 (4.4) |
| Waist hip ratio | 0.9(0.1) | 0.9 (0.1) | 0.9 (0.1) |

## Discussion

This is the first-ever prevalence survey on stroke conducted throughout the country from a representative sample of the population in Bangladesh. This study provides essential information on the prevalence and distribution of common risk factors of stroke in the community.

This survey revealed a high burden of stroke with an overall prevalence of 11.39 per 1000 adult population in Bangladesh, which is higher than other low and middle-income countries (5.36 to 10.40 per thousand), but much lower than the reports (26-80 per thousand) from high-income countries^8-15^. In comparison with our neighboring South Asian countries, the prevalence was similar to the numbers from India; slightly higher than Srilanka but much lower than that in Pakistan (48 per thousand)^9,16,17^. As per global south, the prevalence data from Europe ranges from 14 to 20 per thousand and from different parts of USA range from 18 to 44 per thousand^18,19^.

The prevalence report was nearly 4 times higher than the previously published report from Bangladesh in 2011^20^. This is probably since the previous study was a small-scale survey that involved only 9664 participants from three districts near the Dhaka division. Our data revealed a wide range of geographic variation of prevalence ranging from 7.62/1000 in Rajshahi on the Northwestern part of the country to 14.71/1000 in Mymensingh i.e., north and central part of the country. The finding resonates with various studies from India (9-22/1000), China (6.24-15.49/1000), and the USA (18-44/1000) that involved a nationwide survey of stroke patient^16,18,20,21^.

Quite understandably like the rest of the world, the age-specific prevalence rate showed a predictable rise with the gradual increase of age^18^. It was almost 7 times higher among patients 60 years or older than those younger than 40 years. In line with most of the published studies around the world there was a variation of prevalence across the gender, it was almost two times higher among males (13.62/1000) in this study^9,18^. There were thirteen studies worldwide (India, China, UK, Spain, Italy, USA, Argentina, New Zealand) that measured the gender variation in stroke prevalence across different age groups. The overall prevalence was 41% higher in male^22^. More importantly, multivariate analyses have also indicated that men were two times more likely to develop a stroke than women. The possible explanation might be related to genetic factors and the protective role of estrogen^23^. Similar to the reports from India and China the prevalence was slightly higher in rural areas than the urban areas^21,24,25^. This is probably due to a lack of awareness about the risk factors, changes in dietary pattern, limited access to specialized health care facility, poor control of risk factors, inability to continue medication, etc.

It was not unexpected that we found a major burden of stroke is due to the ischemic subtype. About four-fifth of stroke cases had an ischemic event. This is a common scenario for LMICs^16,26^. In an epidemiologic review from developed countries, Feigin et al showed that up to 67.3–80.5% of strokes are are of ischemic type, whereas 6.5–19.6% are primary intracerebral hemorrhage, about 0.8–7% are subarachnoid hemorrhage, and 2–14.5% are of undefined type^27^.On the other hand, many of the East Asian studies have suggested that the proportion of intracerebral hemorrhage is significantly higher (up to 35% of the total) than in people of European descent^28^. Around two-thirds of the total stroke cases in our survey had at least a CT scan confirmation of stroke which is much higher than the two-year population-based survey conducted in Kolkata that had only half of the patient with a CT scan diagnosis of stroke^16^.

This survey also examined the distribution of the common set of risk factors of stroke. It has been estimated that hypertension accounts for 54% of stroke mortality, followed by dyslipidemia and tobacco smoking in low and middle-income countries^29^. More than three fourth of the stroke cases had hypertension, two-third had the dietary habit of not taking adequate fresh fruits and vegetables, more than a third had dyslipidemia and habit of smoking, around one-fourth had diabetes. A population-based survey (The STEPS survey 2010) in Bangladesh reported that inadequate intake of fruit and vegetables, use of tobacco, low level of physical activity, abdominal obesity, and hypertension were fairly common among the Bangladeshi population^30^. The pool of risk factors we found in this study was similar to the reports from other studies around the world^31^. In consistence with most of the studies around the world, hypertension was the most common factor in our study^32-35^. We also found that dyslipidemia, smoking, and diabetes were three major risk factors among stroke patients. Haque et al. also reported a strong association of LDL-cholesterol and triglyceride with ischemic stroke patients in Bangladesh^36^. In another study, Bak et al. showed the association of smoking in stroke^37^. Regular smoking doubled the risk of stroke among males in Europe, the USA and China^38-42^. We considered smoking and chewable tobacco usage under the same umbrella in this study as smoking is uncommon among Bangladeshi women and they are more habituated to tobacco usage in other forms. The association of stroke with diabetes is well established for the last four decades. A similar finding was also reported in the INTERSTROKE study^43^. Like other epidemiological studies, obesity and ischemic heart disease were also not uncommon in our survey^41,44,45^.

The strength of our study includes, the multistage cluster random sampling technique that involved all the 64 districts throughout the country. The sample size was fairly large enough with a high precision rate in sample size calculation (sampling error of 0.001 and a design effect of 2). All the stroke cases were verified and confirmed by competent neurologists. Still, the survey is not free from it’s limitations. Due to lack of time and funding, we could only report point prevalence. Due to cross sectional design, we could not draw causal inference and evaluate several important risk factors like the contribution of stress, physical activity, etc.

## Conclusion

This nationwide survey yielded a crude prevalence of stroke 11.39 per thousand population. The highest prevalence (14.71/1000) was found in Mymensingh and the lowest in Rajshahi (7.62/1000). The prevalence of stroke in males was almost twice that in females. Rural stroke prevalence was slightly higher. The age-specific prevalence showed a gradual rising trend with increasing age. The stroke prevalence was more than seven times higher among elderly males than their younger counterparts. The majority had an ischemic event. Hypertension, dyslipidemia, tobacco use, diabetes, ischemic heart disease were the most common risk factors observed among stroke patients.

## Data Availability

The database will be available on request.

## Availability of data and materials

The database will be available on request.

## Authors Contribution

BAM was involved in planning the study, setting the methodology for this study. ATM HH was involved in data analysis, data interpretation, and writing the manuscript. The rest of the authors were involved in consultation and data collection. All the authors have read and approved the final version of the manuscript.

## Reference

1. Cohen IK, Fabrizio Fand Bryan M. A simple framework for analyzing the impact of economic growth on non-communicable diseases. Cogent Economics & Finance (2015), 3: 1045215.

2. Feigin VL, Krishnamurthi RV, Parmar P, et al. Update on the Global Burden of Ischemic and Hemorrhagic Stroke in 1990-2013: The GBD 2013 Study. Neuroepidemiology 2015; 45(3):161–76.

3. Feigin VL, Lawes CMM, Bennett DA, Barker-Collo SL, Parag V. Worldwide stroke incidence and early case fatality reported in 56 population-based studies: A systematic review. Lancet Neurol. 2009; 8: 355–69.

4. Bleich SN, Kohelmoos TLP, Rashid M, Peters DH, Anderson G. Non-communicable chronic disease in Bangladesh: Overview of existing programs and priorities going forward. Health Policy. 2011 May; 100(2-3): 282–289.

5. Stroke in Bangladesh [Internet]. World Life Expectancy. 2019 [cited 14 December 2019]. Available from: http://www.Worldlifeexpectancy.com/bangladesh-stroke

6. Islam MN, Moniruzzaman M, Khalil MI, Basri R, Alam MK, Loo KW, and Gan SH. The burden of Stroke in Bangladesh. International Journal of Stroke 2013; 8(3):211–3.

7. Zaman MM, Chowdhury SR, Ahmed J, Akram Hussain SM, MahbubusSobhan SM, Turin TC. Prevalence of stroke in a rural population of Bangladesh. Glob Heart 2015; 10:333–334.

8. Kalkonde YV, Sahane V, Deshmukh MD, Nila S, Mandava P, BangA. High Prevalence of stroke in rural Gadchiroli, India: a community-based study. Neuroepidemiology. 2016; 46:235239.

9. Chang T, Gajasinghe S, and Arambepola C. Prevalence of Stroke and its Risk Factors in Urban Sri Lanka; Population-Based Study. Stroke 2015; 46:2965–68.

10. Khedr EM, Fawi G, Abdela M, Mohammed TA, Ahmed MA, El-FetohNA Zaki AF. Prevalence of ischemic and hemorrhagic strokes in Qena Governorate, Egypt: a community-based study. J Stroke CerebrovascDis. 2014; 23:1843–1848.

11. Fang J, Shaw KM, George MG. Prevalence of stroke -the United States, 2006–2010.MMWR Morb Mortal Wkly Rep. 2012; 61:379–382.

12. de JesúsLlibre J, Valhuerdi A, Fernández O, Llibre JC, Porto R, López AM, Marcheco B, Moreno C. Prevalence of stroke and associated risk factors in older adults in Havana City and Matanzas Provinces, Cuba (10/66 population-based study). MEDICC Rev. 2010; 12:20–26.

13. Truelsen T, Piechowski-Józwiak B, Bonita R, Mathers C, Bogousslavsky J, Boysen G. Stroke incidence and prevalence in Europe: a review of available data. Eur J Neurol. 2006; 13:581–598.

14. Jaillard AS, Hommel M, Mazetti P. Prevalence of stroke at high altitude (3380 m) in Cuzco, a town of Peru. A population-based study. Stroke. 1995; 26:562–568.

15. Aho K, Reunanen A, Aromaa A, Knekt P, Maatela J. Prevalence of stroke in Finland. Stroke. 1986; 17:681–686.

16. Kamalakannan S, Gudlavalleti A, Gudlavalleti V, Goenka S, Kuper H. Incidence & prevalence of stroke in India: A systematic review. Indian Journal of Medical Research. 2017;146 (2):175.

17. Khealani BA, Hameed B, and Mapari UU. Stroke in Pakistan. Journal of the Pakistan Medical Association 2008; 58(7):400–403.

18. Zhang Y, Chapman A, Plested M, Jackson D, Purroy F. The Incidence, Prevalence, and Mortality of Stroke in France, Germany, Italy, Spain, the UK, and the US: A Literature Review. Stroke Research and Treatment. 2012; 2012:1–11.

19. Prevalence of Stroke — United States, 2006–2010 [Internet]. Cdc.gov. 2019 [cited 14 December 2019]. Available from: https://www.cdc.gov/mmwr/preview/mmwrhtml/mm6120a5.htm.

20. Mohammad QD, Habib M, Hoque A, Alam B, Haque B, Hossain S, Rahman KM, and Khan SU. Prevalence of stroke above forty years. Mymensingh Medical Journal 2011; 20 (4): 640–644.

21. Wang W, Jiang B, Sun H, Ru X, Sun D, Wang L, Jiang Y et al. Prevalence, Incidence, and Mortality of Stroke in China: Results from a Nationwide Population-Based Survey of 480 687 Adults. Circulation. 2017; 135:759–771.

22. Peter Appelros, BirgittaStegmayr Andreas Terént. Sex Differences in Stroke Epidemiology: A Systematic Review. Stroke. 2009; 40:1082–1090.

23. Krause, D. N., Duckles, S. P. & Pelligrino, D. A. Influence of sex steroid hormones on cerebrovascular function. J Appl Physiol (1985)101, 1252–1261.

24. Das SK, Banerjee TK, Biswas A, Roy T, Raut DK, Mukherjee CS, et al. A prospective community-based study of stroke in Kolkata, India. Stroke. 2007; 38:906–10.

25. Banerjee TK, Mukherjee CS, Sarkhel A. Stroke in the urban population of Calcutta - An epidemiological study. Neuro epidemiology. 2001; 20:201–7.

26. Oveisgharan S, Sarrafzadegan N, Shirani S, Hosseini S, Hasanzadeh P, Khosravi A. Stroke in Isfahan, Iran: hospital admission and 28-day case fatality rate. Cerebrovasc Dis. 2007; 24(6):495–499.

27. Feigin VL, Lawes CM, Bennett DA, Anderson CS. Stroke epidemiology: a review of population-based studies of incidence, prevalence, and case-fatality in the late 20th century. Lancet Neurol 2003; 2: 43–53.

28. Sudlow CL, Warlow CP. Comparable studies of the incidence of stroke and its pathological types:results from an international collaboration: International Stroke Incidence Collaboration. Stroke 1997; 28: 491–99.

29. Strong K, Mathers C, Bonita R. Preventing stroke: saving lives around the world. Lancet Neurol. 2007; 6:182–187.

30. Zaman MM, Rahman M, Rahman M, Bhuiyan MR, Karim M, Chowdhury MJ. Prevalence of risk factors for non-communicable diseases in Bangladesh: Results from STEPS survey 2010. Indian J Public Health 2016; 60:17–25.

31. Dyken ML, Wolf PA, Barnett HJM, Beran JJ, Hass WK, Kannel WB, Kuller L, Kurtzke YF, Sundt TM. Risk factors in stroke: a statement for physicians by the subcommittee on risk factors and stroke of the stroke council. Stroke. 1984; 15:1105–1111.

32. Al-Rubeaan, K. et al. Ischemic Stroke and Its Risk Factors in a Registry-Based Large Cross-Sectional Diabetic Cohort in a Country Facing a Diabetes Epidemic. J Diabetes Res 2016, 4132589 (2016).

33. Hankey, G. J. et al. Rates and predictors of risk of stroke and its subtypes in diabetes: a prospective observational study. Journal of neurology, neurosurgery, and psychiatry 84, 281–287 (2013).

34. Tseng, C. H., Chong, C. K., Sheu, J. J., Wu, T. H. & Tseng, C.P. Prevalence and risk factors for stroke in Type 2 diabetic patients in Taiwan: a cross-sectional survey of a national sample by telephone interview. Diabetic medicine: a journal of the British Diabetic Association 22, 477–482 (2005).

35. Guerrero-Romero, F. & Rodriguez-Moran, M. Proteinuria is an independent risk factor for ischemic stroke in non-insulindependent diabetes mellitus. Stroke; a journal of cerebral circulation 30, 1787–1791 (1999).

36. Haque Akmf, Islam QT, Rahman KM et al. Serum Lipid as a Risk Factor of Ischaemic Stroke in Bangladeshi People. J MEDICINE 2012; 13:22–26.

37. Bak S, Bak L, Sorense JS. Prevalence of risk factors in cerebral ischemia. Ugeskr Laeger 1995; Jan 23; 157:444–6.

38. Asplund K, Karvanen J, Giampaoli S et al. Relative Risks for Stroke by Age, Sex, and Population Basedon Follow-Up of 18 European Populations in the MORGAM Project. Stroke. 2009; 40:2319–2326.

39. Rodriguez BL, D’Agostino R, Abbott RD, Kagan A, Burchfiel CM, Yano K, Ross GW, Silbershatz H, Higgins MW, Popper J, Wolf PA, Curb JD. Risk of hospitalized stroke in men enrolled in the Honolulu Heart Program and the Framingham study: a comparison of incidence and risk factor effects. Stroke. 2002; 33:230 –236.

40. Curb JD, Abbott RD, MacLean CJ, Rodriguez BL, Burchfiel CM, Sharp DS, Ross GW, Yano K. Age-related changes in stroke risk in men with hypertension and normal blood pressure. Stroke. 1996; 27:819–824.

41. Jorgensen HS, Nakayama H, Raaschou HO, et al. Intracerebral hemorrhage versus infarction: stroke severity, risk factors and prognosis. Ann Neurol 1995; 38:45–50.

42. Zhang XF, Attia J, D’Este C, Yu XH, Wu XG. A risk score predicted coronary heart disease and stroke in a Chinese cohort. J Clin Epidemiol. 2005; 58:951–958.

43. O’Donnell MJ, Xavier D, Liu L, Zhang H, Chin SL, Melacini PR et al. Risk factors for ischaemic and intracerebral haemorrhagic stroke in 22 countries (the INTERSTROKE study): a case-control study. Lancet 2010: 376; 112–23

44. Jood K, Jern C, Wilhelmsen L, Rosengren A. Body mass index in mid-life is associated with a first stroke in men: a prospective population study over 28 years. Stroke. 2004; 35:2764 –2769.

45. Tanne D, Medalie JH, Goldbourt U. Body fat distribution and long-term risk of stroke mortality. Stroke. 2005; 36:1021–1025.

